# Epidemiology study of SARS-CoV-2 pandemic in India, the first and second wave

**DOI:** 10.1101/2021.12.20.21268016

**Authors:** Dwaipayan Chaudhuri, Joyeeta Datta, Satyabrata Majumder, Kalyan Giri

## Abstract

**Background and objectives:** SARS-CoV-2 has wrecked the world for the past 17 months. India has been hit by the second wave of the virus which has been characterized by new symptoms. This study focuses on the pattern of infection over the last 13 months utilizing epidemic model to predict course of the pandemic.

**Material and methods:** The data was collected from covid19india.org to perform analysis based on age and gender distribution. Statistical analysis was performed to determine the relation between confirmed and recovered cases while SIR epidemic model was used to determine the course of the pandemic in the country and the changes that have occurred from the first to the second wave.

**Results and discussions:** Results show infectivity rate to be higher in ages 20-50 while mortality is higher in 50-80 age group while 60-70% of the infected population are males. Each of the 9 states have their own salient feature curves of infection. It was seen that the confirmed and recovered cases are more correlated at present than previous wave. The curves for both waves show a polynomial distribution while the reproduction number data shows an almost U-shaped curve indicating decrease of infection spread in the middle phase when the first wave was on a decline before picking up again owing to the second wave.

**Interpretations and conclusion:** The gender and age distribution shows that although lower age group is more infected, mortality is high for higher age groups, on the other hand males are more prone to the infection. The statistical analysis shows the nature of spread of the disease, the data of which is quantified by the SIR model based study.

## Introduction

Coronavirus disease 2019 i.e. COVID-19 has turned into a global pandemic which is continuing till date in several countries including India [1]. The median age of patients affected by COVID-19 has been found to be 47 years [5]. While the country is going through the devastating second wave of virus infection, the unprecedented surge in confirmed cases could be a result of several different factors including the emergence of particularly infectious novel variants, a rise in unrestricted social interactions, and low vaccine coverage [2].

With the ever-growing curve of confirmed and recovered cases in case of the SARS-CoV-2 infection, our study takes a look into the gender and age distributions of the infection. The study not only considers India as a whole but also focuses on 9 states where the virus has accounted for most number of reported cases. The study also focuses on the correlation between the confirmed and recovered cases and makes use of the SIR epidemic model to calculate the infection and recovery rate in various phases of the pandemic in India and finally the reproduction number is calculated. This study may give us insight on the spread of the virus in the country and give a view on where we stand in this pandemic.

## Materials and methods

### 1. Epidemiology based analysis

The daily infectivity data of SARS-CoV-2 was taken from the open database covid19india.org till 20th April 2021. Analysis was performed taking into consideration gender and age distribution data of all infected people till August, 2020 after which only deceased people data were taken, since data for the entire infected category was not available. Data was analyzed on a state by state basis that include Andhra Pradesh, Delhi, Karnataka, Kerala, Maharashtra, Rajasthan, Tamil Nadu, West Bengal and Uttar Pradesh.

### 2. Statistical and Mathematical analysis

Correlation and regression analysis of daily confirmed and recovered cases were performed to determine the correlation between the two. The SIR epidemiology model was applied to calculate the infection rate, recovery rate and reproduction number based on data from March, 2020 to 20th April, 2021 [3,4].

In the exponential curve equation, y = e^rx^, Gamma (γ) is average of recovery rate over the period, Beta (β = e^r^ + γ − 1) is calculated from the equation of exponential growth and SIR model differential equations. R_o_ (R_o_= β/γ) is the reproduction number which means number of people infected by single infected person. R_o_ denotes the average number of secondary infections caused by an infected host.

## Result and Discussions

### 1. Epidemiology based analysis

Over the course of the last 13 months it has been seen that the daily active cases curve gradually increased, reached a peak around mid-September to October after which the curve gradually decreased. But since end of February, 2021, it has been seen that the curve started to rise and the tremendous increase led to the more devastating second wave of the SARS-CoV-2 pandemic. For the recovered cases, a balance was maintained in the first wave, but in case of the second wave the balance has been completely destroyed since the increase in cases happened dramatically. In case of October, 2020 to February, 2021, when the first wave was dying down it was seen that the difference between daily confirmed cases and daily recovery even sunk to below 0 giving negative results indicating that the recovery rate has surpassed the infection rate. But for the last month it has been seen that this difference has surged to new heights.

The daily death and recovery rates indicate that over the course of time the mortality rate has decreased dramatically from hovering around 3-4% coming down to below 1% at present. Besides this the recovery rate increased dramatically, where between October, 2020 to February, 2021 it was constantly above 100% with the value even reaching as high as 180% but due to the surge in number of cases it has come down to below 60% although it has started rising again.

The positivity rate which reached a highest of around 35% during the peak of first wave sank to below 5% in the first part of the year but that percentage has as well grown to reach almost 45% and even surpassing it which makes the second wave very dangerous since less number of confirmed cases were found before in spite of large number of RT PCR tests being performed but in this scenario the positivity rate has increased by almost 10 times in a matter of days.

In case of the first 5 months when the complete data was available and even after that, the data of the deceased cases, it was seen that 60-70% cases were of males. This data proves that the males are much more prone to the disease than females.

In case of age distribution, it was seen that for the confirmed cases in the first 5 months, the majority of the affected people belonged to the age group of 20-50 while for the latter time period when only data of the deceased were seen it was observed that most of the deceased people belonged to the age group from 50 to 80 and more specifically 60 to 80. This shows that although most of the affected people were younger but the mortality rate was much higher in case of the elderly population than the younger population.

The state by state analysis for 9 states where the maximum number of confirmed SARS-CoV-2 cases has been reported till 20th April, 2021 (Table 1).

**Fig 1:**
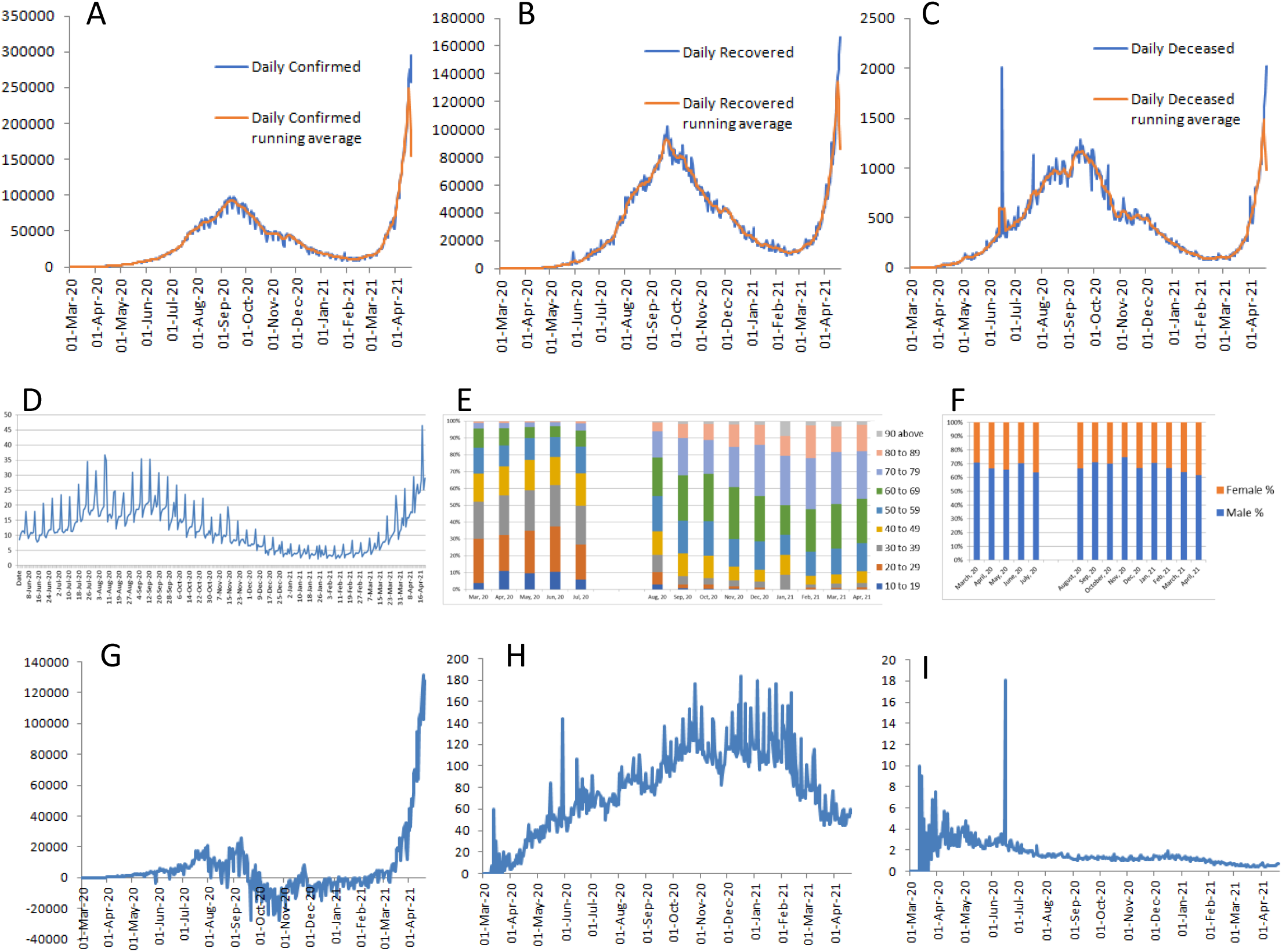
Epidemiology based analysis of cases in India. Monthwise raw data and running average curves for A. Confirmed, B. Recovered, C. Deceased cases, D. Positive rate in India, E. Age distribution of confirmed and deceased cases, F. Gender distribution of confirmed and deceased cases, G. Difference between daily confirmed and recovered cases, H. Daily death ratio, I. Daily recovery rate.

**Fig 2:**
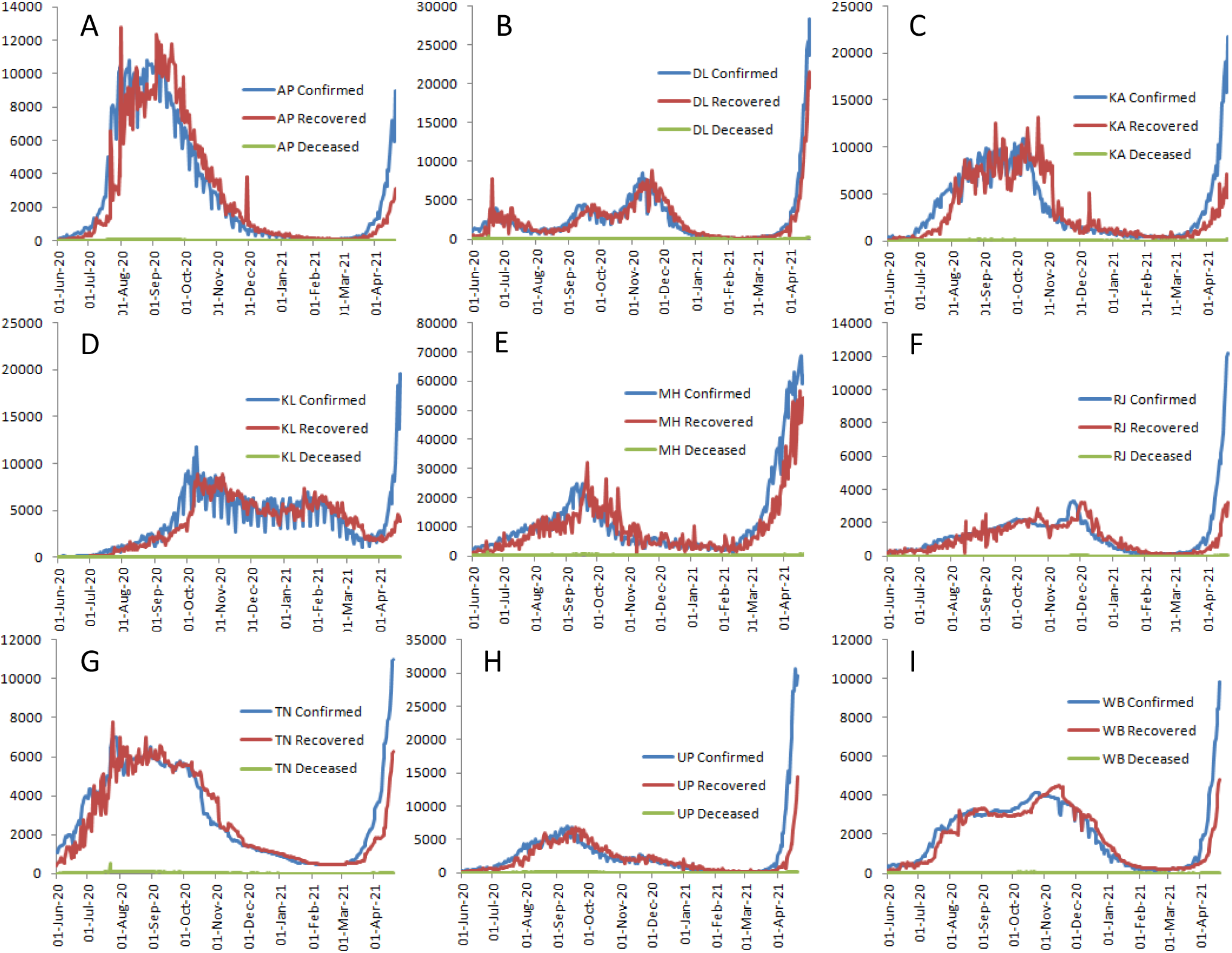
state wise Case graph. A. Andhra Pradesh, B. Delhi, C. Karnataka, D. Kerala, E. Maharashtra, F. Rajasthan, G. Tamil Nadu, H. Uttar Pradesh, I. West Bengal

**Table 1:** State-wise data of first and second wave

| State | First wave | Second wave | Comments |
| --- | --- | --- | --- |
| Andhra Pradesh | July to August-September, 2020 | March, 2021 onwards | Difference between confirmed and recovered cases has become wide unlike first wave |
| Delhi | mid-June, mid-September to mid-November, 2020 | March, 2021 onwards | 4 spikes have been seen with height of peak increasing with each wave<br>Recovery rate in the second wave has not dropped alarmingly similar to the previous 3 peaks |
| Karnataka | July to December, 2020, peak around September-October, 2020 | March, 2021 onwards | Recovery curve lagged behind the confirmed case curve<br>Second wave peak is already twice the height of first wave peak leading to an increase of difference between daily confirmed and recovered cases |
| Kerala | August, 2020 to February, 2021 | March, 2021 onwards | Second wave is a continuation of the first wave as there was no particular trough period |
| Maharashtra | peak around mid-September, 2020 | March, 2021 onwards | In the first wave almost perfect normal distribution of confirmed cases seen<br>October onwards recovered cases surpassed daily confirmed cases<br>In the last month steep rise in both confirmed cases and recovered cases seen |
| Rajasthan | September, 2020 to January, 2021 | late February, 2021 onwards | The first wave was seen as a plateau with no particular peak being found<br>In most states the infectivity rate dropped in the late 2020 and early 2021 but here the trend was not prominently observed |
| Tamil Nadu | cases started to increase in July, 2020; peaked near August where it turned into a plateau and was maintained till October | March, 2021 onwards | In the second wave a steep increase of both daily confirmed cases and daily recovered cases seen |
| Uttar Pradesh | August to November, 2020 | March, 2021 onwards | low level of infection was seen even after first wave passed for 2 months following which the daily confirmed cases almost reached the base of the line i.e almost 0 |
| West Bengal | mid-July to mid-December | end of March, 2021 | In spite of plateau being formed in the first wave without any prominent peak, the caseload increased around mid-October, 2020; second wave curve has increased steeply |

### 2. Statistical analysis

Based on regression analysis it was seen that over the course of one year extending from March 2020 to March 2021, the regression line between daily confirmed cases and daily recovered cases had an equation of y = 0.95x + 473.77. The same line when data from April 2020 to April 2021 was taken into account was seen to be y = 0.63x + 8913.41. The data for first and second wave was seen to be a quadratic type.

**Fig 3:**
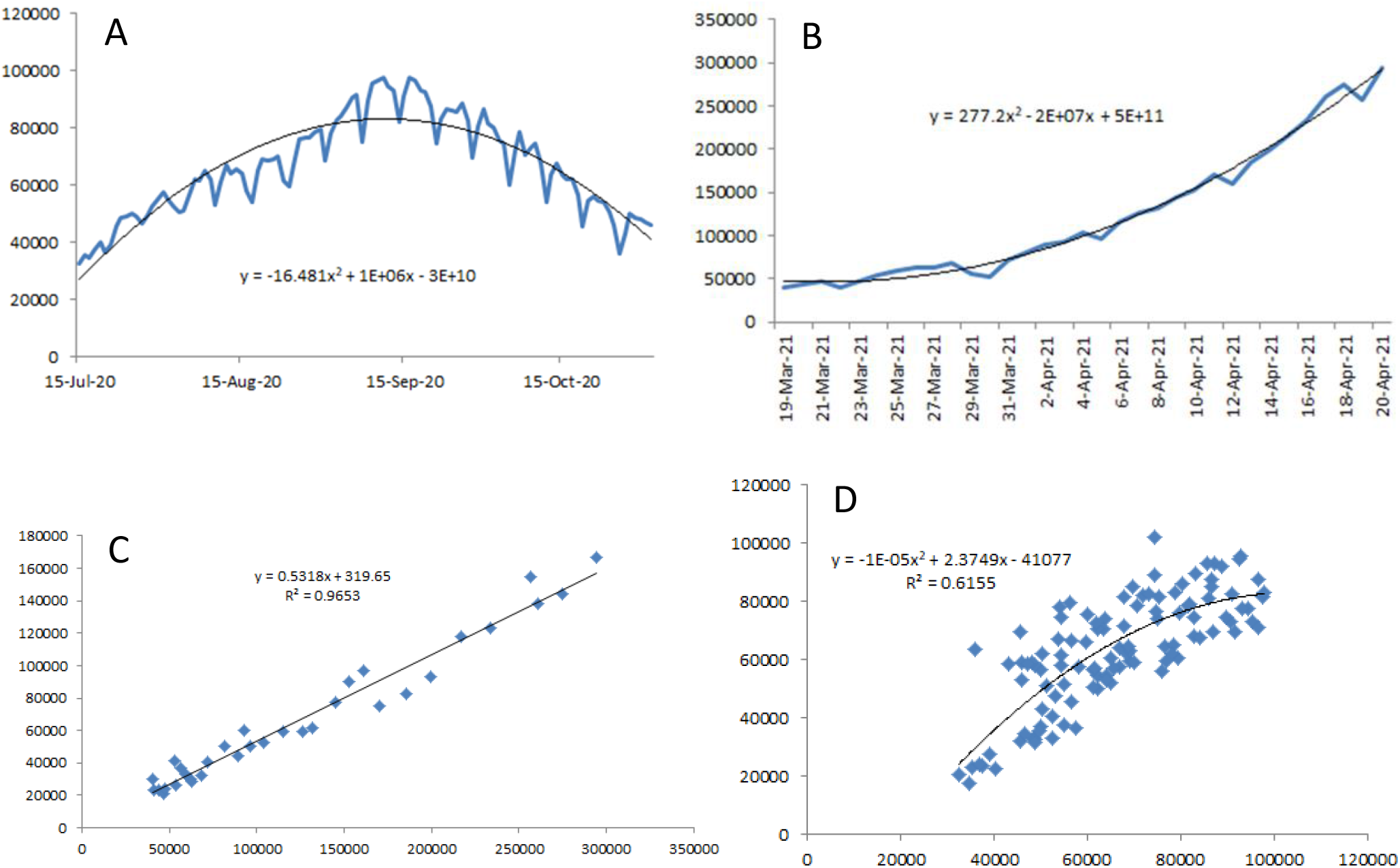
Statistical analysis of SARS-CoV-2 cases in India. A. Curve and equation of curve for first wave of infection from 15th July, 2020 to 1st November, 2020, B. Curve and equation of curve for second wave of infection from 15th March, 2021 to 20th April, 2021, C. Regression line for second wave of infection between daily confirmed and recovered cases, D. Regression line for first wave of infection between daily confirmed and recovered cases

The Pearson correlation coefficient (R^2^) for March 2020-2021 was seen to be 0.91 thus showing almost perfect positive correlation between confirmed cases and recovery but the value dropped to 0.88 when the time range was from April 2020-2021 and when data was taken for 1 year from mid-April 2020-2021 it is 0.78. The value is also evident from the graph where in April 2021, where an increase of difference between the two curves is observed. The month by month change in correlation between confirmed and recovered cases is provided in Table 2.

**Table 2:** Month wise SIR model data for infection rate, recovery rate, reproduction number and R^2^.

| Month | Beta- infection rate | Gamma-recovery rate | reproduction number | $R^2$ |
| --- | --- | --- | --- | --- |
| April, 20 | 0.27 | 0.22 | 1.23 | 0.86 |
| May, 20 | 0.54 | 0.5 | 1.08 | 0.54 |
| June, 20 | 0.66 | 0.63 | 1.05 | 0.8 |
| July, 20 | 0.72 | 0.68 | 1.06 | 0.92 |
| August, 20 | 0.89 | 0.88 | 1.01 | 0.47 |
| September, 20 | 0.92 | 0.93 | 0.98 | 0.001 |
| October, 20 | 1.19 | 1.21 | 0.98 | 0.65 |
| November, 20 | 1.1 | 1.11 | 0.99 | 0.2 |
| December, 20 | 1.19 | 1.22 | 0.97 | 0.72 |
| January, 21 | 1.17 | 1.19 | 0.98 | 0.4 |
| February, 21 | 1.05 | 1.03 | 1.02 | 0.08 |
| March, 21 | 0.74 | 0.68 | 1.1 | 0.79 |
| April, 21 | 0.61 | 0.53 | 1.15 | 0.94 |

### 3. Mathematical model based analysis

SIR model was used to calculate beta, gamma and R_0_. Based on the curve of daily confirmed cases, the curve has been divided into 5 regions. For each of the regions, beta, gamma and R_0_ were calculated to see the change in virus infectivity and extent of recovery in the patients in each of these regions.

**Fig 4:**
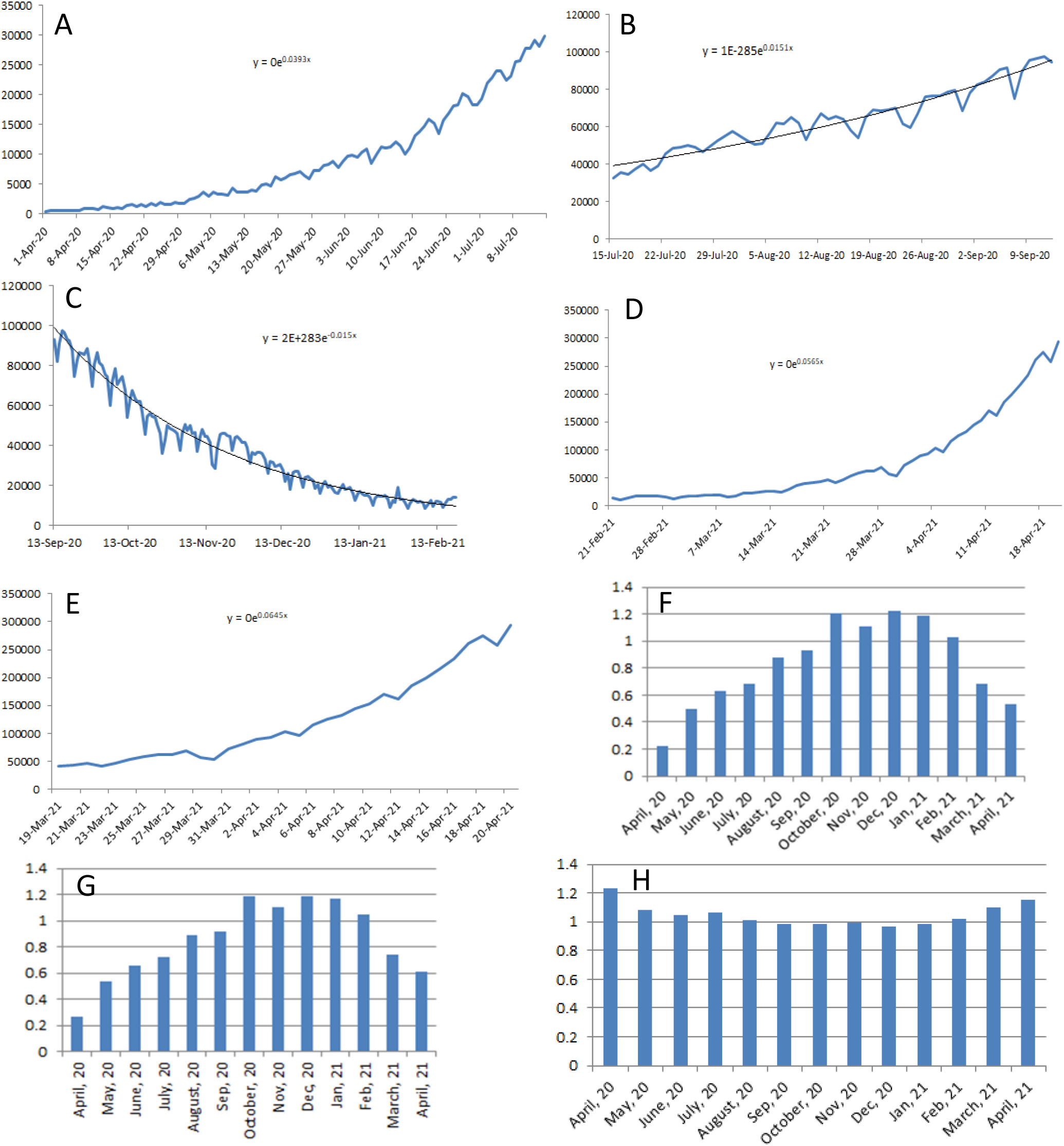
SIR model data deduced from confirmed and recovered cases in India. The first 5 panels from A to E correspond to the exponential curves according to the date ranges as follows: A. from 1st April, 2020 to 14th July, 2020, B. from 15th July, 2020 to 12th September, 2020, C. from 13th September, 2020 to 20th February, 2021, D. from 21st February, 2021 to 20th April, 2021, E. from 19th March, 2021 to 20th April, 2021, F. month wise recovery rate (gamma), G. month wise infection rate (beta), H. month wise reproduction number.

Based on month by month analysis it has been seen that the reproduction number, was below 1 from September, 2020 to January, 2021, time when the first wave curve started to fall. It has to be seen that for the picking up of both the waves the R_o_ value did not exceed 1.2 value. For the infection rate and recovery rate, it was seen that the rate gradually increased and reached a peak and then started to decrease with the maximum of both curves in October 2020, December 2020 and January 2021. If the R_o_ value is greater than one, the infection rate is greater than the recovery rate, and thus the infection will grow throughout the population. If R_o_ is less than one, the infection quickly will die out since people are healing faster than they are spreading it. Thus it can be said that the infection is in growth phase of the second wave at the moment.

## Conclusion

This study shows the change in nature from first wave of infection to the second wave. It has been seen that the younger generation is more infected while older generation have a higher mortality with the males being predominantly infected. The data suggest an increase in infection and recovery rate, thus keeping the reproduction number under control till now. The reproduction number of 1.2 is the maximum that it has attained. It is also seen that the confirmed and recovered cases weren’t very much correlated during the first wave but in the second the daily data is highly positively correlated as seen from regression analysis. This study would help to determine the future course of infection by extrapolating the data over future time range.

## Data Availability

All data produced are available online at covid19india.org

https://www.covid19india.org/

## Notes

### Competing Interest Statement

The authors have declared no competing interest.

### Funding Statement

This study did not receive any funding

### Author Declarations

covid19india.org

